# Community-Delivered Antenatal Care to Address Access Barriers: Perspectives of Women and Healthcare Workers

**DOI:** 10.64898/2026.09.23.26363848

**Authors:** Olive Kabajaasi, Winfred Nakato Nansozi, Ashley Younger, Judith Kiconco, Shanitah Nankya, Daniel Lukakamwa, Assen Kamwesigye, Ibrahim Bwaga, Andrew Odur, Rose Mukisa-Bisoborwa, Wessel Ganzevoort, Sanne Gordijn, Kerstin Klipstein-Grobusch, Marcus Rijken, Josaphat Byamugisha, Aris T. Papageorghiou, Sam Ali

## Abstract

Stillbirth remains a major public health concern, particularly in resource-constrained settings. Barriers to timely, high-quality antenatal care may delay the recognition and management of pregnancy complications. We explored the perspectives of women, Village Health Team (VHT) members and healthcare workers on community-delivered approaches to improve pregnancy care, in particular prevention of stillbirth in Uganda.

An exploratory qualitative study was conducted at four referral hospitals in Uganda. Data were collected through focus group discussions, in-depth interviews, and key informants’ interviews with women, Village Health Team members and healthcare workers. Audio-recordings were transcribed verbatim and translated into English where required. Data were analysed thematically using an iteratively developed codebook and NVivo software.

In total, 56 participants took part, comprising women (n=27), Village Health Team members (n=11) and frontline healthcare workers (n=18). Participants highlighted the need for the adoption of simple and user-friendly wearable pregnancy monitoring tools for self-monitoring of early warning signs. Health education sessions focused on identifying early warning signs of pregnancy complications highlighted, emphasizing the importance of empowering women with information to seek care early through digital platforms. Limited follow-up support was also a critical concern, with the majority of the women advocating for better emotional and psychological care after a stillbirth, both at a facility and at home.

Participants viewed community-delivered antenatal care as an acceptable approach to reducing access barriers. Implementation would require trained personnel, quality-assured tools, clear referral pathways and integration with facility-based care. Structured communication and psychosocial support after stillbirth were also identified as priorities. These findings should inform co-design and prospective evaluation of community-delivered models.

## INTRODUCTION

Stillbirth, classified as the death of a fetus after 28 weeks of gestation, before or during birth, remains a major global health concern. Low-income countries (LMICs) bear the greatest burden with Sub-Saharan Africa accounting for the highest proportion worldwide [1,]. Stillbirth has profound psychological, social, and economic consequences for women and their families and is associated with grief, depression, financial hardship, stigma and social isolation [2].Recognizing this burden, the “Every Newborn Action Plan (ENAP)”, adopted by the World Health Assembly in 2014, set a global target of 12 or fewer third-trimester stillbirths per 1,000 total deliveries by 2030. Progress remains insufficient and projections suggest that nearly 7.7 million babies may be stillborn in Sub-Saharan Africa by 2030, with countries not achieving the ENAP target without intensifying interventions [3].

Stillbirths are associated with a range of maternal, placental and fetal causes and risk factors including post-term pregnancy, maternal infections such as malaria, syphilis, HIV, hypertension, diabetes, and fetal growth restriction [4],[5]. Additional contributors include intrapartum complications, fetal asphyxia, trauma, and congenital infection [6]. Although not all stillbirths are preventable, timely, high-quality antenatal care (ANC) can identify and manage several associated conditions. The World Health Organization (WHO) therefore emphasizes improving the quality of antenatal care as a key strategy to reduce stillbirth through early detection and management of pregnancy complications, improved access to care, effective triage and timely referral from pregnancy to safe delivery [7].

In Uganda, several strategies have been implemented to address known risk factors for stillbirth, including scaling up basic and comprehensive emergency obstetric care services, periconceptional folic acid supplementation, malaria prevention and improved detection and management of syphilis [8]. Nevertheless, stillbirth rates remain unacceptably high [9], [10].

The WHO recommends initiation of ANC within the first 12 weeks of gestation, and a total of eight contacts during pregnancy [7]–[11]. Evidence from Uganda highlights persistent barriers to early and consistent ANC utilization. A study conducted in Mulago National Referral Hospital has shown that delays in reaching referral facilities significantly increase the risk of stillbirth [7], [8]. Additional barriers include limited paternal involvement, perceived poor quality of care, health care system constraints, socio-cultural beliefs, fear of HIV testing and long distances to health facilities [14][9]. These barriers contribute to delayed care-seeking, missed opportunities for early detection of pregnancy complications, and ultimately increased stillbirth risk.

Recent technological advances offer opportunities for early detection and prediction of pregnancy complications [10]. For example, studies have demonstrated that the use of an electronic fetal heart rate monitor can improve the women’s birth experience [15]–[17]. Similarly, the Ultrasonic Cardiac Output Monitor (USCOM) has shown to be, a non-invasive, reliable and user-friendly tool for assessing maternal cardiovascular function in pregnancy [11]. Previous work has highlighted the importance of embedding ultrasound examination for biometric measurements and vascular resistance within clinical practice guidelines and ensuring adequate training of healthcare providers to prevent inappropriate clinical decisions arising from misinterpretation of ultrasound or Doppler findings [12][13]. While facility-based technological interventions are promising, their uptake and effectiveness may be limited by health systems constraints and access to care barriers particularly in low-resource settings. Consequently, community-based approaches for early screening, risk identification and timely referral may be critical to improving stillbirth prevention. This study therefore aimed to explore stakeholders’ perspectives on early detection and prevention of stillbirth in Uganda.

This qualitative study was nested within a larger multi-site prospective clinical observational study (iTECH) which sought to develop and validate risk-prediction models for pregnancy complications associated with stillbirth, including pre-eclampsia, fetal growth restriction, and Gestational Diabetes Mellitus, using maternal characteristics, factors, liquid biomarkers, hemodynamics measures, and ultrasound markers[14]. Umbilical cord blood and placenta tissues samples were collected for selected cases and controls, and processed for subsequent analysis. The qualitative component of this study was reported in accordance with the Consolidated Criteria for Reporting Qualitative Research (COREQ) guidelines [15].

## METHODS

### Study design

This study employed an exploratory qualitative design to understand perspectives on early detection and prevention of stillbirth. Data were prospectively collected from 02 July, 2024 to 27 September 2024 using focus group discussions (FGDs), in-depth interviews (IDIs) and key informants’ interviews (KIIs) with key stakeholders. Including women, Village Heath Team members (VHTs) and healthcare workers. This approach enabled an in-depth exploration of lived experiences, perceptions, and contextual factors influencing antenatal care access, early risk identification, and stillbirth prevention.

### Study setting

The study was conducted across four large public study sites implementing the iTECH study in Uganda: Kawempe National Referral Hospital (KNRH) in the central region, Hoima National Referral Hospital (HRRH) in the western region, Lira Regional Referral Hospital (LRRH) in the northern region, and Mbale Regional Referral Hospital (MRRH) in the eastern Region. Sites were purposively selected to ensure capture variation in geography, population and health system context. The hospitals differed in patient volume, staffing, and annual delivery capacity (table 1).

**Table 1:** Site demographics.

| Site | Total bed capacity | Annual admissions | Annual deliveries |
| --- | --- | --- | --- |
| Kawempe NRH | 381-bed women's and children hospital | 60,814 | 18,634 births |
| Hoima RRH | 317-bed capacity | 27,099 | 7,308 births |
| Lira RRH | 400-beds | 28,958 | 6,028 births |
| Mbale RRH | 250 beds (52 maternity beds) | 58,169 | 7,816 births |

*Source: 2023/2024: Uganda Ministry of Health Annual Performance Sector Report [16].*

### Participant selection and recruitment

Participants were selected using both purposive and convenience sampling techniques guided by the principle of maximum variation to ensure representation across the different stakeholder groups. The sample comprised pregnant women, non-pregnant women with prior stillbirth, non-pregnant women with a positive pregnancy outcome, Village Health Team members, and healthcare workers involved in antenatal and maternity care. Pregnant women and VHT members were recruited prospectively according to availability and relevant characteristics, while women with prior pregnancy outcomes were recruited retrospectively based on time since outcome, age, and parity. Initial contact was made by iTECH community engagement officers via telephone to assess willingness to participate. Those who agreed were invited to the health facility, provided with detailed study information, and gave written informed consent prior to participation. Healthcare workers were recruited through face-to-face engagement at the facilities. Women were eligible if they were aged 18 years or older, enrolled in iTECH, attending antenatal care at participating sites, and had experienced a stillbirth or a positive pregnancy outcome at least one month before the interview. Healthcare workers were eligible if they were involved in obstetric, midwifery, or antenatal services and had experience caring for pregnant women or managing stillbirths. Healthcare workers were eligible if they were involved in obstetric, midwifery, or antenatal services and had experience caring for pregnant women or managing stillbirths. Women awaiting delivery or cesarean section, those experiencing acute grief, and healthcare workers not involved in antenatal care were excluded.

### Sample size

Participants were recruited across several stakeholder groups. Four FGDs were conducted with pregnant women (one at each site) and two with VHT members; all other participants completed individual interviews. Of 81 people approached, 56 participated. The six FGDs included 31 participants (20 pregnant women and 11 VHT members). Individual interviews were conducted with four women who had experienced stillbirth, three women with a positive pregnancy outcome and 18 healthcare workers, including three hospital managers interviewed as key informants. Recruitment continued until the study team judged that no substantively new themes were emerging. Reasons for non-participation included lack of partner permission, time constraints, loss to follow-up and lack of interest. The recruitment approaches, data collection methods, and number of participants included in each participant category are summarized in Table 2 below.

**Table 2:** Recruitment and sampling methods.

| Participant category | Recruitment | Data-collection method | Number of participants |
| --- | --- | --- | --- |
| Pregnant women receiving ANC | Convenience sampling | Focus group discussions (4) | 20 |
| Women with previous stillbirth | Purposive sampling | In-depth interviews | 4 |
| Women with a positive pregnancy outcome | Purposive sampling | In-depth interviews | 3 |
| Healthcare workers in ANC clinics and iTECH | Purposive sampling | Semi-structured interviews | 15 |
| Hospital managers (Obstetricians) | Purposive sampling | Key informant interviews | 3 |
| Village Health Team members | Convenience sampling | Focus group discussions (2) | 11 |
| **Total** |  |  | **56** |

### Data Collection

Between July and September 2024, data were collected by a trained social scientist (OK) with experience in qualitative health research, assisted by a note taker (AY). Data collection methods included semi-structured IDIs, FGS, and KII’s, using study guides jointly developed by the study team. The guides consisted of open-ended questions designed to facilitate clear and comfortable participant responses (S1 Appendix FGD Guides and S2 Appendix Interview guides). Interviews and FGDs were conducted in private rooms within hospital settings and were audio-recorded with participant consent. Sessions were conducted in English, Luganda, Lugisu, Langi, or Runyoro based on participant preference. FGDs lasted 42 to 76 minutes, while individual interviews ranged from 24 to 69 minutes. No enrolled participants withdrew from the study. Following interviews at the first two study sites (Kawempe and Hoima), the research team held debriefing discussions to reflect on emerging findings and assess data saturation. Initial themes and codes were documented in a notebook; the study team took no interview-specific field notes.

### Data Analysis

Audio-recordings were transcribed verbatim by an independent consultant and translated into English where required. OK checked each transcript against the recording for accuracy. OK and AY independently read each transcript to familiarize themselves with the data, noted preliminary themes and jointly developed an initial codebook. Transcripts were not returned to participants for comment. OK led coding in NVivo 12 (QST International, Cambridge, MA) by and AY reviewed the coding and verified the analysis in NVivo 14. The codebook and themes were refined through iterative discussions with the study team until consensus was reached. Transcripts were not returned to participants for comment.

### Ethics approval and consent to participate

This qualitative study was conducted as part of the approved iTECH project and received ethical clearance by the Makerere University School of Medicine (SOMREC; Mak-SOMREC-2022-535) and the Uganda National Council for Science and Technology (UNCST; HS2762ES) as documented in supporting files (S4 IRB Approval letter and S5 UNCST Approval letter). The study compiled with ethical standards as outlined in the Belmont Report. All participants provided written informed consent after receiving information about the study procedures and confidentiality safeguards. Participants received approximately 8USD as compensation for their time and related inconveniences. Participation in the study was voluntary and participants were free to withdraw at any time without penalty. To ensure confidentiality, audio recordings were anonymised using unique identification codes instead of names. Audio files were destroyed after transcription.

## RESULTS

Participants included 18 healthcare workers of different cadres (4 midwives performing USCOM and arteriography, 4 sonographers, 1 research assistant, 3 obstetricians and 4 field officers). The sample also included 20 pregnant women, four women who experienced a stillbirth, three women with positive pregnancy outcomes, and 11 VHTs. Half of the women interviewed had previously lost a pregnancy, and their parity ranged from one to six children. Healthcare workers’ experience ranged from two to twenty years and over. Female participants constituted 45 (80%), healthcare workers 18 (27%), and local leaders 11 (19.6%). The participant age range was 18-65 years. The characteristics of participants with in respective categories are presented in table 3 below.

**Table 3.**
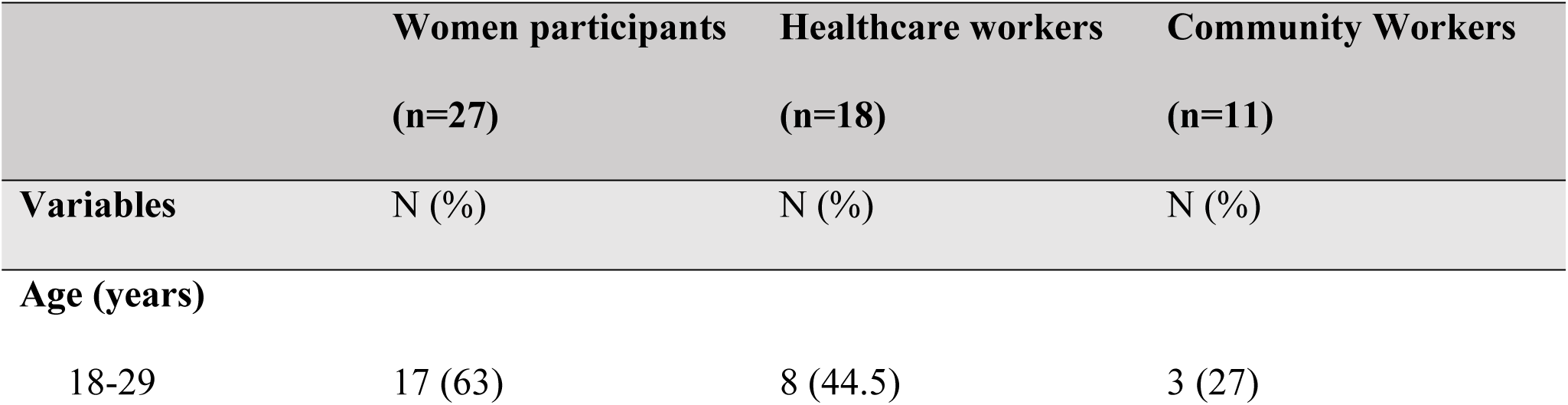
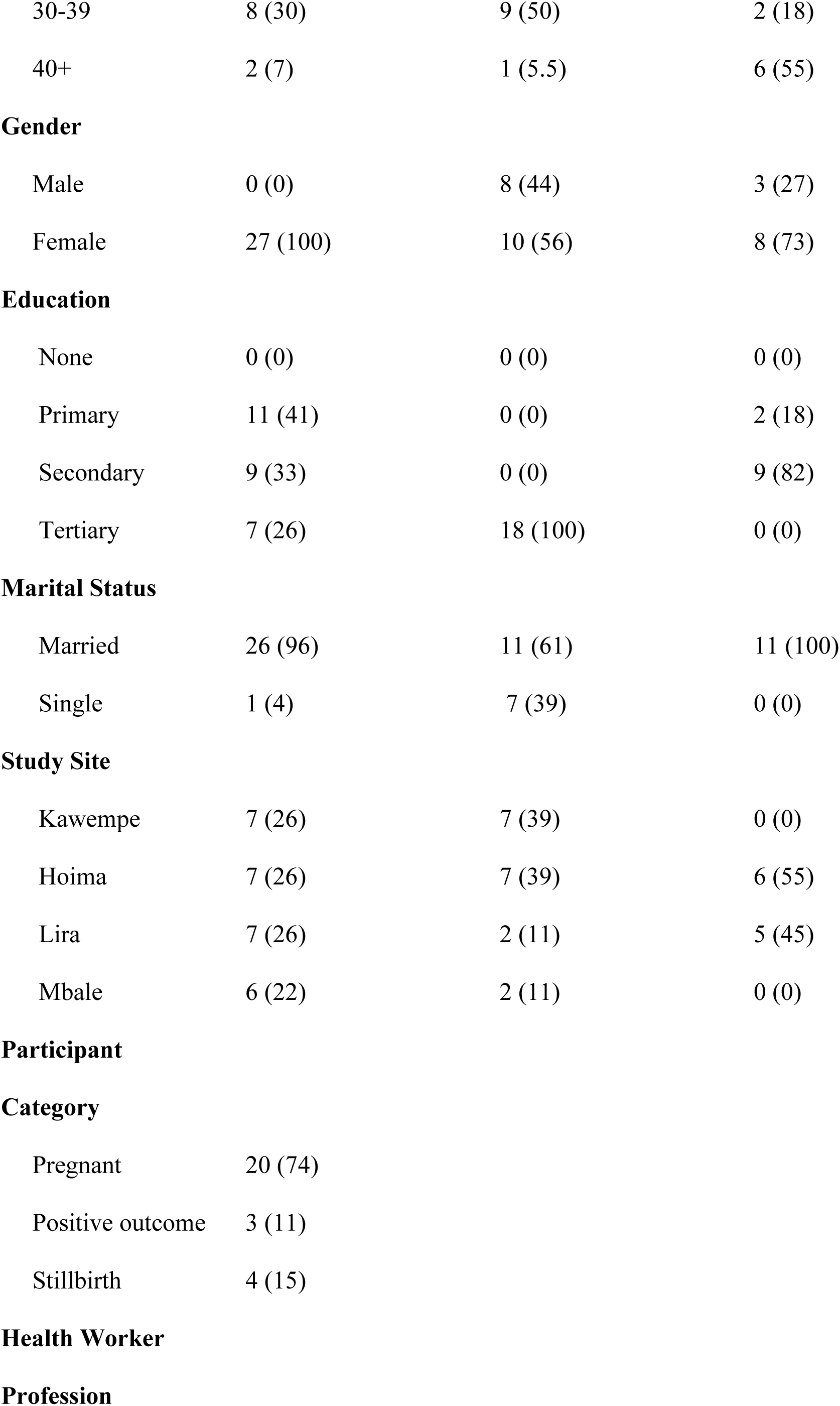
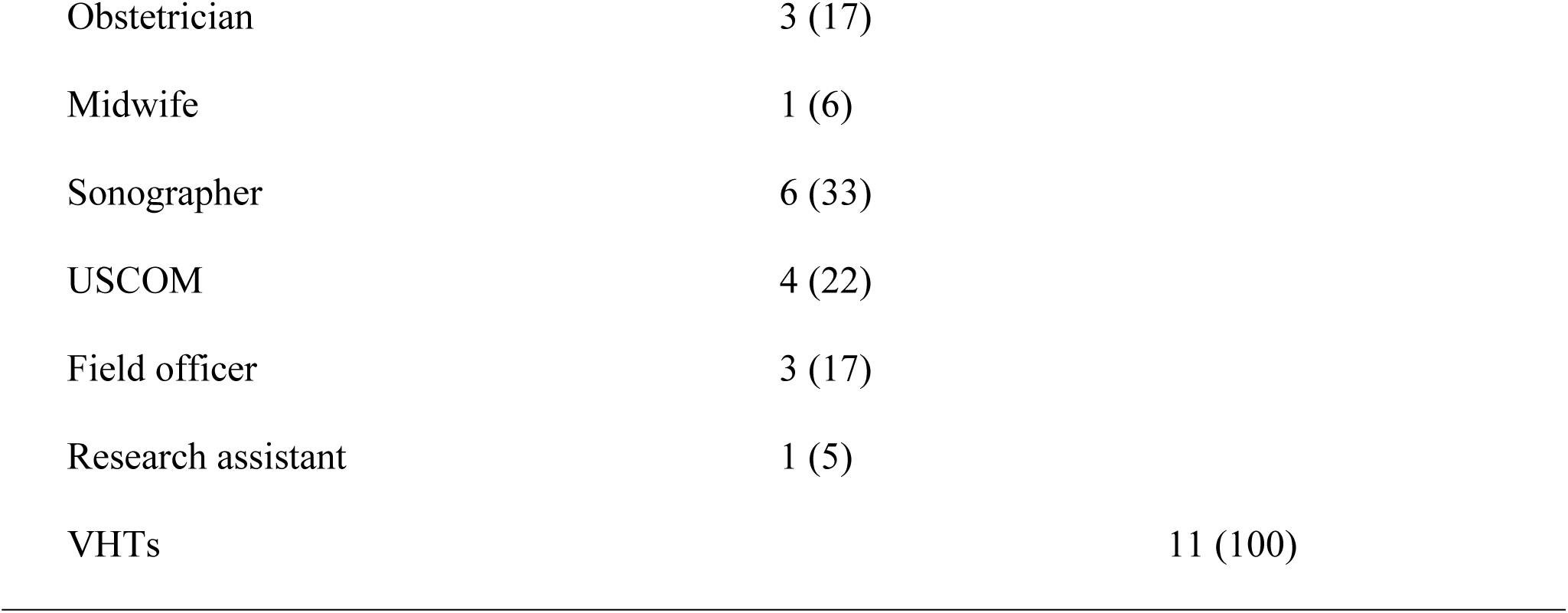
Characteristics of participants within respective categories.

In this study, we identified two overarching, broad themes: 1) Community based monitoring and 2) Follow-up care. These, and the identified five sub-themes, and seven child nodes are presented in table 4 below.

**Table 4:**
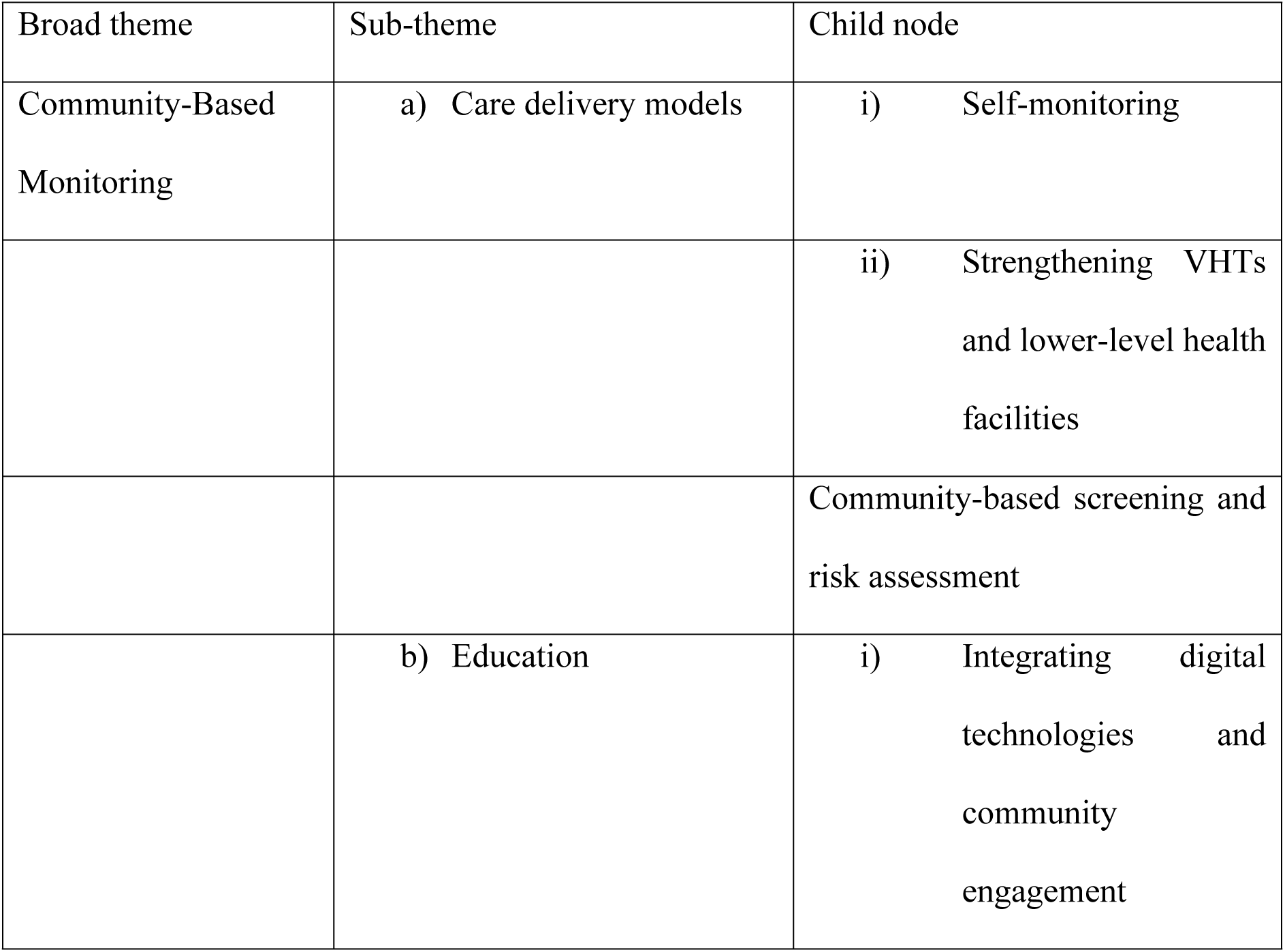
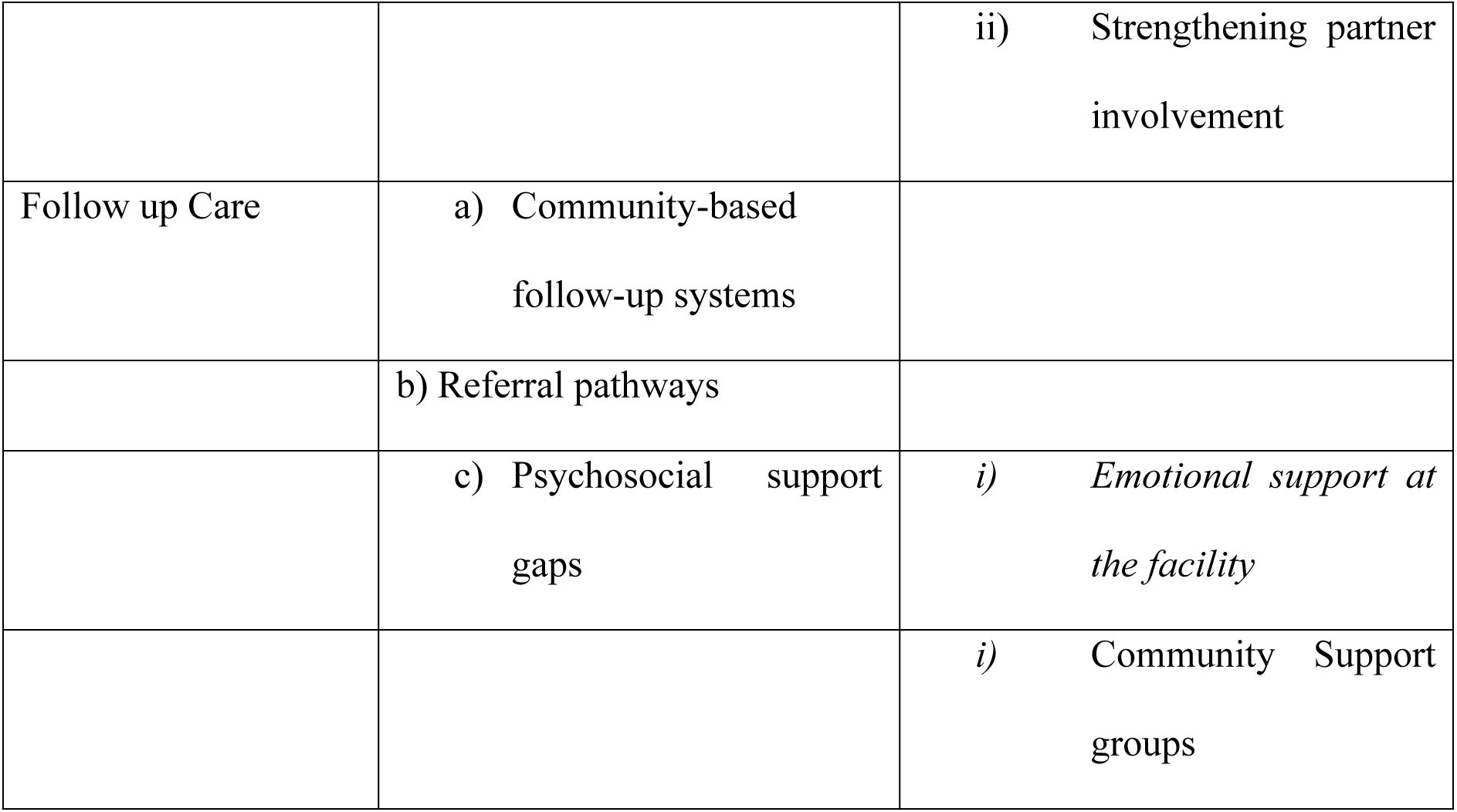
Themes, sub-themes and child nodes.

## COMMUNITY BASED MONITORING

This theme covers two sub-themes: care delivery models and education as explained in detail below.

### Care delivery models

#### Self-monitoring

The first delivery model proposed by most participants was at-home monitoring to guide women to make care seeking decisions. Participants preferred simple and user-friendly wearable pregnancy monitoring tools that they can use to monitor early warning signs at their convenience. HCWs wanted pregnant women to carry out self-tests for high blood pressure, diabetes, bleeding and to track foetal movements that can directly be linked to the health facilities for urgent care:

#### Strengthening VHTs and lower-level health facilities

One proposed care delivery model involves empowering VHTs and lower-level health facilities to provide essential maternal health services that would otherwise necessitate travel to distant higher-level healthcare facilities. Participants, especially women, stressed that VHTs are community members and have the potential to foster trust and uptake of services, especially in rural areas.

> “You can reach out to these mothers through their leaders, the VHTs because they believe in their VHTs and even Local Councils and if these are the very people who are educating them. They will quickly grasp the information that these people are giving them. Instead of you the health workers coming from nowhere and you’re teaching them. They will not even listen to what you’re saying but if you pass through their leaders, the VHTs, their chair persons and counsellors and their religious leaders. These people trust these people as if they are gods, mostly the pastors, whatever their leaders say, they go with it. Whatever the priests say they go with it. So if you can go through these leaders, it will be a walk over. You will not hear them being rebellions at all. HCW Hoima.
>
> “We have our VHTs, they can test us from the community, they have been delivering malaria drugs for the children and we always report to them. They usually educate us on other diseases but not pregnancy. They can test us very well”. Woman who had a positive outcome.

In addition, participants were of the view that VHTs can be empowered to conduct pregnancy-related tests under supervision from healthcare providers to help healthcare workers to remain focused on more urgent and complex pregnancy issues. Health workers at the lower health facilities in particular added that this model would work well by training health workers, and providing them with the necessary essential tools would be essential.

> “So if the lower health facilities are supported… because most of our mothers start from health center two or three then come here. So if these (lower) facilities are supported with machines and weighing scales, the health workers are given maybe some efficiency training maybe on basic objective care they will improve on the diagnosis of conditions that lead to stillbirths. Like you find a mother has come, she has not taken any blood pressure and she is in the third trimester and has come for labour, no result of blood pressure but she has been attending antenatal care. Or a mother has come and she has been getting half of the package she is supposed to get. So, the mother thinks she has been going to the health facility but you realize the services she has been getting is incomplete. So the problem may come and she goes not attended to and ends into stillbirth. So if you can support these facilities at the lower level. It would be very good”. Male HCW, Lira.

#### Community-based screening and risk assessment

Apart from self-monitoring, participants had strong support for community-based screening for pregnancy complications conducted by trained HCWs through community outreaches, to bring pregnancy services closer to home and solve some of the financial challenges related to pregnancy. They were of the view that simple tests such as blood pressure, HIV, malaria screening, blood work and pregnancy scans should be conducted within the communities.

> “Women in the community don’t have money to go the hospital. So the scans should be taken to them in the community because when you go to a health center you find there is no scan and you also find that there is no money to travel. They can get a very small scan which they can just pass on the mother’s tummy and if there is a pregnancy she will know, if she had blood pressure she will know, if she has a fever she will know”. Pregnant woman Lira FGD.
>
> “Other assessments can be done in the communities like once in a while like you can bump into a community. Like you can say this time we are going into such a community and when you go there do that. There is some health facility where I went and they were doing it like family planning; they were giving them from the community. They would get a clean room, they put their instruments and they do the work. Then they could palpate mothers from there and when they realize the mother is pregnant, they give her the next appointment like when you come to the hospital and they were coming. Yes, they were testing and then others were palpating and if they realize that you’re pregnant and you never started antenatal, they link you to a health facility for the next appointment. So I don’t know if that one also can be done to other communities”. Female HCW, Mbale.

### Education

Education was the second component of community-based monitoring. Health education programs that focus on recognizing early warning signs of pregnancy complications were identified, emphasizing the importance of empowering women with information to seek care early. Under this theme, two sub-topics were identified: integrating digital technologies and community engagement as well as strengthening partner involvement:

#### Integrating digital technologies and community engagement

Study participants suggested the use of mobile technology as a useful tool in providing information about pregnancy care. Participants talked about adopting easy-to-access digital media platforms through which information could be shared. These included: WhatsApp, radio, and television. Local methods included face-to-face health talks, smaller group meetings and one-on-one meetings on pregnancy to support women in understanding pregnancy care, adopting healthy behaviour, and identifying early warning signs of stillbirth. In the quote below, a participant highlights the different forms of education and also emphasizes the importance of involving local leaders in disseminating this information:

> “Now days we use social media, and WhatsApp, because we are urbanized. But when we go to the rural areas, it completely becomes a challenge, they do not have TVs but they have radios, which they use batteries that can go for a month. If possible, the radio talk shows can be supplemented with health talks in the village, we invite chairman to call upon people and the community so that we talk to them. That would have helped us, information would spread quickly in that, the one who attended the meeting would inform those who did not attend. So, that was my request”. Female VHT member.

#### Strengthening partner involvement

Participants frequently described limited partner support during ANC, which often leaves women to handle pregnancy-related tasks alone, causing physical and emotional challenges. Women in particular emphasized the need for male involvement in pregnancy, given that men make decisions regarding healthcare access. To reduce this gap, participants recommended appropriate education programs targeting male partners including HCWs being in direct contact with male partners, to improve support for women.

> “I put my husband’s number there, even this time iTECH, told him, for me to come, but when you tell him that you’re going to hospital, he will ask what you are going to do there? Weren’t you at the same hospital last time? So, in order for me not to argue, I put his number there, so they could be calling him. He called me and told me that, they said on Friday, they need you at the hospital, in fact, he is the one who woke me up even today.”-Pregnant women FGD, Kawempe.
>
> “Information regarding the causes of stillbirths and how they can be avoided, plus the male involvement, should always be there. Emphasizing involvement like the so called financial providers and they run in everything plus the male figures. But again, the man is the head of the family so he should be there to make decisions that will impact the family including where the woman should seek care. Not only to provide transport.”-HCW Kawempe

### FOLLOW-UP CARE

#### Community-based follow-up systems

Women and healthcare providers reported a lack of appropriate follow-up structures to support women in accessing the needed care, be it before or even after experiencing complications. HCWs and women reported that only women who experienced stillbirth mentioned iTECH’s approach, which involved a telephone call and a home visit to provide education and emotional support to the families after experiencing stillbirth. They found this support helpful when expressing and sharing their grief. However, this was not the same with women who did not participate in the iTECH project.

> *‘It is very good, because the doctor wants you to know the truth, was it from the baby or from the mother, that’s it. Because the baby might be in the womb but when they have their issues like the baby or sometimes, it is the mother who has caused the problem”. “They explain everything, they give you time and even ask you to come back to the hospital if you aren’t feeling well. They are very friendly”.* Woman who experienced a stillbirth.

#### Referral pathways

Participants reported that the health facilities and the communities were not well linked to support appropriate referrals to the hospitals. HCW talked about how they are not able to connect with pregnant women requiring emergency obstetric care to health facilities. This lack of communication affects HCWs ability to prepare for high-risk cases. They also noted the challenge of out-of-stock essential supplies that further delays timely care. To address these challenges, participants suggested leveraging the existing referral standards to ensure women are easily directed to seek care.

> “So when it comes to having anti-susceptive supplies, cotton, gloves, all these, this is now hospital and we run out of supplies like I said because when NMS supplies, within two weeks’ things are done. The sutures get done within less than two weeks. So when the patient is due for C-section for example, and the hospital doesn’t have, at such times patients and caretakers are told to avail…but if you delay to intervene and you ask for fluids and they are not in place, the patient has to buy. So if she can’t afford eventually this prolonged distress might now result into the affixture and below score now four”. Obstetrician.

### Psychosocial support gaps

#### Emotional support at the facility

Women who lost their pregnancies reported feeling angry and blamed themselves, while also expressing that HCWs appeared too busy and only focused on treating the medical symptoms, with less explanation or mental support. Women also reported sharing rooms with other women who had live babies, which intensified their grief and anger, highlighting the need for strong psychosocial support both at the facility and at home. Here is what one participant shared.

> “I was not even understanding myself, even if you tapped me, I wouldn’t even hear you, my thoughts were very far (deep in thoughts). They didn’t even explain anything to me; I wanted to know what killed my baby but I didn’t get the answer”. Woman who experienced a Stillbirth, Lira.

#### Community support groups

HCWs strongly highlighted the need for support groups both at the facility and in the community to promote sharing, learning, and help women cope with the loss. They noted that while they provide immediate medical care, they are not able to meet all the women’s emotional and social needs; therefore, support groups are needed to offer ongoing support to women.

> “So they should be creating support groups like those mothers who lose their babies they can be incorporated into and they make support groups. So that when they are in those groups everyone shares their challenges and how they are coping. So it will help these mothers so that they know that I am not the only one experiencing this. Because now I see these support groups are helping these HIV positive people. They help these mothers to adhere to their treatment and suppress their viral load and l see forming these support groups will help these mothers to move on. They will know they are not the only one with this problem there are other people who are experiencing what am experiencing but now are okay and they have been able to deliver other babies and are living a happy life”. Female HCW Mbale.

## DISCUSSION

This study sought to assess perceptions of women and HCWs on prevention and prediction of stillbirth in Uganda. Participants highlighted the need for community-delivered ANC models to help them independently monitor and track their pregnancies using simple and user-friendly digital tools. This could improve early detection and timely intervention of pregnancy-related complications, and aligns with a growing body of evidence on self-monitoring as a way to enhance patient engagement and autonomy. Limited follow-up support intervention was a critical concern, with the majority of the women advocating for better emotional and psychological care after a stillbirth, both at the facility and at home. They also recommended health education sessions focused on identifying early warning signs of pregnancy complications, emphasizing the importance of empowering women with information to seek care early through digital platforms.

Studies have shown that self-monitoring of BP during pregnancy can lead to earlier identification of hypertensive disorders, prompting faster medical attention and reducing adverse outcomes [17]. Similarly, self-monitoring of blood glucose has been associated with improved glycemic control among pregnant women with gestational diabetes [18]. By empowering women with technology-driven self-monitoring tools, gaps in accessing facility-based maternal care, particularly in resource-limited settings where regular clinic visits may be challenging, could be reduced. However, while participants recognized the benefits of this approach, their perspectives also underscore key challenges that must be addressed, such as affordability, user training, devices reliability, results interpretation and timely response to abnormal results. In light of these challenges, prior studies have highlighted the need for clear guidelines for healthcare workers (HCWs)and timely response to abnormal readings [19]. These self-monitoring safeguards are important as community-based monitoring could lead to increased anxiety among pregnant women, which may worsen the already existing pregnancy condition or even further delay care-seeking.

Our findings further show the need for a holistic approach to maternal care by prioritizing not only the medical but also the emotional and psychological well-being of women. Participants recognized the importance of psychosocial support both in the community and at the health facility. This form of support not only reduces stress but also provides opportunities for women to share their pregnancy-related experiences and feel cared for. At the facility, psychosocial care can help patients feel more heard and supported, which often leads to better engagement with treatment and follow-up care. Studies have shown that when people feel emotionally supported, they are more likely to stick to their treatment plans and report better overall well-being. Research has shown that one in ten pregnant women experience anxiety disorder related to low income or bad relationships [20]. Future interventions may need to consider appropriate referral mechanisms to reduce the stress and helplessness that may often come with high-risk pregnancies. There is a need to incorporate counseling, mental health screenings, and appropriate communication into routine care to improve women’s experiences and health outcomes. In addition, training healthcare workers on psychosocial support and integrating peer support groups or community-based mental health interventions could also help bridge this gap. The absence of psychosocial support not only affected women’s mental well-being but also influenced their confidence in the healthcare system. Some women reported feeling unsupported and less likely to urgently report to the hospital unless the symptoms are severe, making it necessary to design appropriate follow up strategies for women.

Our findings also underscore the importance of Community Health Workers in strengthening referrals and linkage. Community Health Workers such as Village Health Team members play a critical role in bridging the gap between the community and the healthcare system, yet they often lack the training or resources to support follow-up care effectively. The findings are consistent with a study in Northern Uganda, which highlighted that the role of VHTs could be a significant means of achieving universal primary healthcare; however, these lacked motivation, transport, and adequate skills [21][22]. HCWs are already stretched, with multiple roles and responsibilities. Without the support of community volunteers, they may lose track of which women need follow-up. Investing in training, incentives, and integrating the VHT services into formal health services could significantly improve follow-up care for women who might receive abnormal screening results in the community. More so, training healthcare workers to provide appropriate counseling and building community-based peer support groups could help ensure that women feel safe and supported through their pregnancy periods.

## Strengths and Limitations

This study has several strengths. First, it was conducted across multiple sites, capturing perspectives from women and health workers in four major regions in Uganda, which helped to ensure that the study findings are not limited to one geographical area. Second, the study included a wide range of stakeholders, including pregnant women, those with normal deliveries, and those who experienced a stillbirth, which provided a comprehensive understanding of the different experiences and perceptions surrounding pregnancy and stillbirth. Limitations that should be considered are that participants were convenience sampling used for some groups; their views may differ from those of women who do not access formal antenatal care or live further from referral services. Recall and social-desirability bias are possible, particularly because some healthcare workers were involved in the iTECH study, but as these staff also provide routine care we believe this is less likely. Finally, participant preferences cannot establish feasibility, diagnostic accuracy, safety, cost-effectiveness or impact on stillbirth. The proposed models require co-design and prospective evaluation.

## CONCLUSION

This study highlights the need for improved pregnancy care models that extend beyond health facilities and respond to women’s needs. Women and healthcare workers emphasized the importance of community-based monitoring approaches, including self-monitoring, and outreach screening, as practical ways to improve early detection of pregnancy complications and reduce barriers to care. While community-based follow-up care, referral systems, and emotional support were perceived as beneficial when available, these services remain fragmented and inconsistently implemented. Addressing these gaps through community facility linkages, referral pathways, and psychosocial support, including support groups for women who experience stillbirths, has the potential to improve women’s experiences of care, support recovery after loss, and contribute to better maternal and perinatal outcomes.

## Data Availability

The datasets used and/or analysed during the current study are available from the corresponding author on reasonable request.

## Acknowledgements

We would like to thank the women, healthcare workers and management at the study sites for providing invaluable information about the study topic.

## Author contributions

OK and AY carried out data analysis and drafted the manuscript with regular inputs from SA. OK, WNN, AY, RMB, WG, SG, KKG, MR, JB, ATP, and SA contributed to study design, data collection methods and tools. OK, carried out the interviews. WNN, AY, JC, SN, DL, AK, IB, AO contributed to site data collection activities. All the authors critically reviewed the work for intellectual content, and approved the final manuscript.

